# Prevalence and factors associated with prediabetes and undiagnosed diabetes in Cambodia: Cross-sectional study based on the World Health Survey Plus 2023

**DOI:** 10.1101/2025.03.09.25322832

**Authors:** Srean Chhim, Grace Marie Ku, Paul Kowal, Vannarath Te, Mony Rithisey, Chamnab Ngor, Poppy Walton, Khin Thiri Maung, Nawi Ng, Por Ir, Kerstin Klipstein-Grobusch, Chhorvann Chhea, Heng Sopheab

**Author notes:** ***Corresponding author:*** Srean Chhim (#109, street 01, Krang Thnong, Saensokh, Phnom Penh, postal 120804).

## Abstract

**Background:** This study aimed to determine the prevalence and factors associated with pre-type 2 diabetes (pre-T2D) and undiagnosed T2D (UDD) in Cambodia.

**Methods:** This cross-sectional study included 5,271 individuals aged ≥ 18 years from all provinces in Cambodia. Pre-T2D and UDD were defined using the World Health Organization (WHO)’s haemoglobin A1c criteria.

**Results:** The weighted prevalences of pre-T2D and UDD were 27.2% and 10.9%. Pre-T2D prevalence was higher in urban areas compared to rural areas (aOR = 1.2, 95% CI: 1.0 - 1.4), females aged 30-39 (aOR=1.4, 95% CI: 1.0 - 2.0), females 40-49 (aOR=2.4, 95% CI: 1.7 - 3.3), females 50+ (aOR = 3.8, 95% CI: 2.8 - 5.1), and males aged 30-39 (aOR=2.0, 95% CI: 1.3 - 3.0), males 40-49 (aOR=2.3, 95% CI: 1.5 - 3.3), males 50+ (aOR = 3.3, 95% CI: 2.4 - 4.7) relative to females aged 18-29, overweight individuals (aOR = 1.6, 95% CI: 1.3 - 1.9), obese individuals (aOR = 1.9, 95% CI: 1.5 - 2.4), those with high waist circumference (aOR = 1.5, 95% CI: 1.2 - 1.8), and elevated total triglycerides (aOR = 1.3, 95% CI: 1.1 - 1.5). Similar risk factors were identified for UDD, with the addition of elevated blood pressure (aOR = 1.5, 95% CI: 1.1 - 2.1).

**Conclusion:** The high prevalence of pre-T2D and UDD in Cambodia is a pressing public health concern. Urgent and intensive interventions are needed to effectively prevent and manage T2D, particularly among urban residents, older adults, and individuals with metabolic risk factors.

**Key messages:** *What is already known on this topic:*

- In 2023, the national prevalence of pre-type 2 diabetes (pre-T2D), measured by impaired fasting glycaemia, among adults (18+) in Cambodia was estimated to be 5.5%, with a higher prevalence in older adults and females.
- The prevalence of undiagnosed T2D (UDD) was not estimated.

*What this study adds:*

- This study revealed a significant prevalence of pre-T2D (27.2%) and UDD (10.9%) among adults (18+ years old) in Cambodia, indicating a substantial public health challenge that requires immediate attention.
- This study identified modifiable and non-modifiable factors associated with pre-T2D and UDD, including urban residence, older age, overweight or obese, high waist circumference, elevated triglyceride levels, and elevated blood pressure.

*How this study might affect research, practice, or policy:*

- These findings underscore the importance of prevention and screening initiatives aimed at early detection to mitigate T2D and to delay or prevent complications in individuals with UDD.

## INTRODUCTION

Type 2 diabetes (T2D) continues to be a leading disease burden and represents a significant global public health challenge.^1^ Adding to this challenge are populations with pre-type 2 diabetes (pre-T2D) that have blood sugar levels higher than normal but not high enough to be diagnosed with T2D.^2^ Specialised institutions in T2D prevention and management like the World Health Organization (WHO), American Diabetes Association (ADA), and International Expert Committee (IEC), define pre-T2D inconsistently, with at least five definitions identified: (1) ADA: fasting blood glucose (FBG) 100-125 mg/dl, (2) WHO: FBG 110-125 mg/dl, (3) ADA/WHO: 2-h post-load blood glucose (2hBG) 140-199 mg/dl, (4) WHO: HbA1c 5.7-6.4%, and (5) IEC: HbA1c 6.0-6.4%.^2^

Up to 70% of individuals with pre-T2D may progress to T2D, with the estimated prevalence increasing exponentially.^3^ The global prevalence of pre-T2D, as measured by the ADA’s FBG criteria, was 9.1% (464 million) in 2021 and is projected to increase to 10.0% (638 million) by 2045.^4^ The prevalence of pre-T2D tended to be higher when measured using HbA1c.^5–8^ Furthermore, approximately half of the global population with T2D remains unaware that they have T2D, increasing the probability of complications and impedes effective T2D management. ^9^ Certain risk factors for T2D, such as age, sex, genetics, ethnicity, history of gestational diabetes, delivery of large-for-gestational-age infants, and menopause, are non-modifiable, and addressing modifiable risk factors can mitigate the risk of developing pre-T2D and T2D.^10^ ^15^ Modifiable risk factors include unhealthy dietary habits, physical inactivity, tobacco use, excessive alcohol consumption, poor stress management, and poor sleep patterns, which are also risk factors for metabolic syndrome and a constellation of intermediate risks, including elevated blood pressure and abnormal cholesterol/triglyceride levels.^10^ ^11^

In Cambodia, the prevention, screening, and management of T2D face challenges within the pluralistic healthcare system. One study assessed key components of integrated T2D care and found a low implementation score in early detection, primary care treatment, health education, self-management support, structured collaboration, and care organisation.^12^ Furthermore, only 10% of general outpatient visits and 35% of T2D-specific visits occurred in public facilities, indicating that most visits were made to private sectors where patients were required to pay out-of-pocket expenses. ^13^ ^14^

Research on pre-T2D and UDD in Cambodia is limited. In 2023, a nationally representative non-communicable disease (NCD) risk factor survey using FBG examined the prevalence of T2D and its associated risk factors (physical inactivity, alcohol consumption, tobacco use, sodium intake, diet, and high blood pressure).^15^ This study estimated pre-T2D prevalence at 6.3% but did not estimate the prevalence of UDD.^15^ Another study in Cambodia assessed the proportion and risk factors of UDD but focused on individuals aged 40 years and older.^16^ Importantly, the study excluded younger adult populations who may also be at risk.^16^

To address these gaps, our study aimed to contribute to understanding T2D in Cambodia by examining the prevalence of, and factors associated with, pre-T2D and UDD. These results provide valuable insights into the development of effective T2D prevention and management strategies in Cambodia.

### Conceptual framework

The prevalence and factors associated with pre-T2D and UDD have been extensively studied in other settings, including Southeast Asia.^17–23^ Pre-T2D and UDD typically have a mix of non-modifiable and modifiable risk factors (Figure 1).^24^

**Figure 1.**
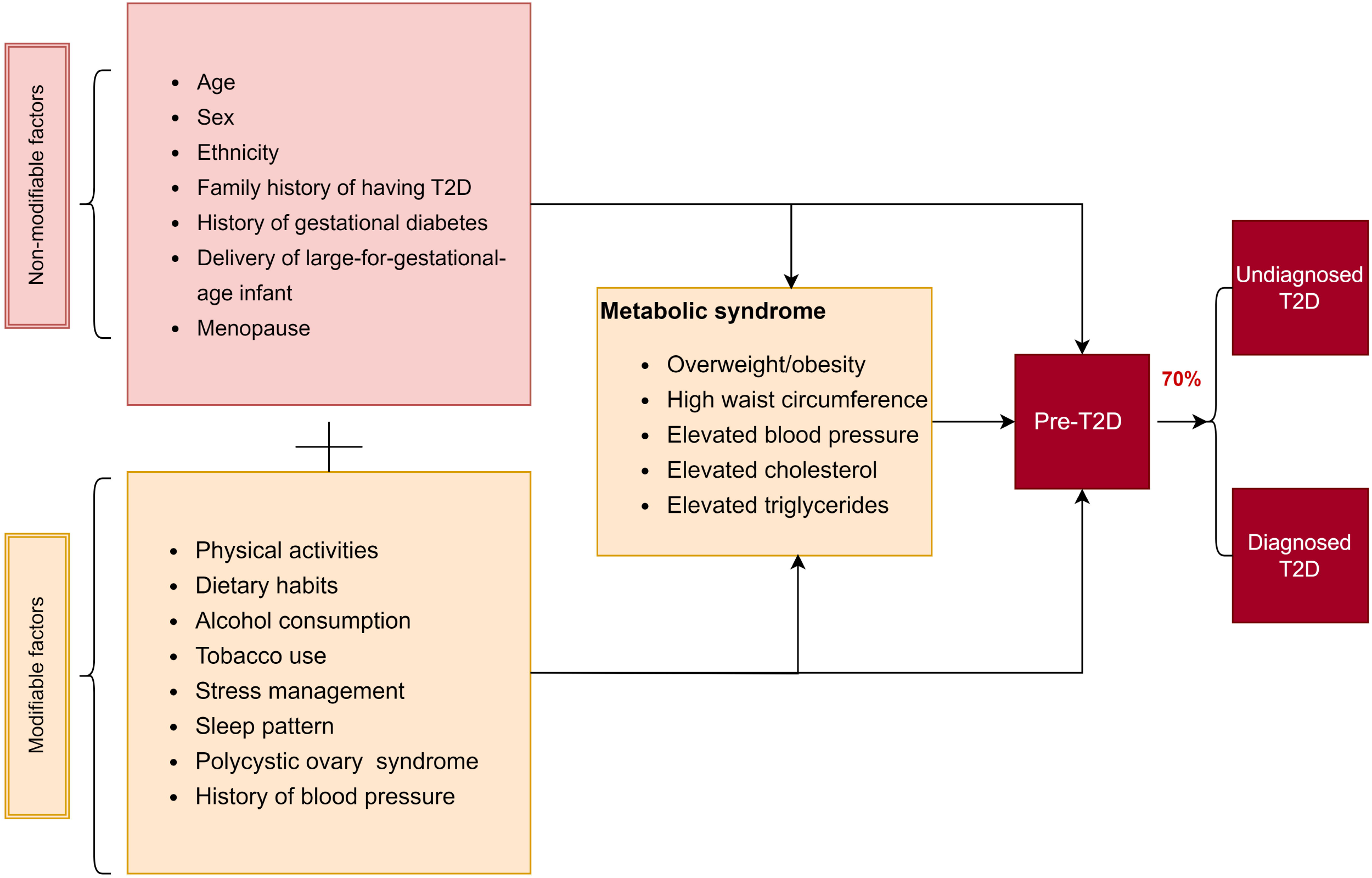
Theoretical framework of the potential association between risk factors and pre-T2D and UDD.

Previous studies have shown that age is an established independent factor, as human cells become resistant to insulin or the pancreas produces insufficient insulin with advancing age. Age can also be associated with reduced physical activity, leading to weight gain and an increased risk of pre-T2D and T2D. Physical activity and other factors, including dietary habits, alcohol consumption, tobacco use, stress management, and sleep habits, can be interrelated and strongly associated with metabolic syndrome before developing T2D.^25–27^ The analysis of pre-T2D and UDD factors should consider a multifaceted approach that includes non-modifiable and modifiable factors. This approach can help identify high-risk individuals and inform targeted interventions for early detection and management of T2D.

## METHODS

### Data sources

This nationally representative cross-sectional study used data from Cambodia’s 2023 World Health Survey Plus (WHS+).^28^ The WHS+ is a WHO-led global tool tracking progress towards health-related goals in low-and middle-income countries.^28^ The National Institute of Public Health in Cambodia, WHO, HelpAge International, and the University of Gothenburg in Sweden conducted the survey in Cambodia. It involved interviews with a nationally representative sample of adults aged ≥ 18 years from selected households nationwide.

### Sampling and recruitment process

A three-stage sampling process was used for the WHS+. First, 276 of the 14,568 villages were randomly chosen across Cambodia, with a selection probability proportional to the village size. Second, we selected households. High-resolution satellite images and GIS software were used to map all buildings within each selected village. From this, 44 buildings were randomly selected based on the pre-testing data, indicating that approximately 50% would contain eligible households. Households were eligible if they had at least one permanent resident aged 18 years or older who had lived there for at least six months in the last 12 months (n=6,154). Within each household, one eligible adult was randomly selected for an individual interview, and the household head completed a separate household questionnaire. Of 6,154 eligible households, 5,271 completed the survey, yielding a response rate of 85.6%. For the prevalence calculation, 844 participants were excluded owing to unavailable HbA1c results (Supplementary Figure 1).

### Data Collection

Data were collected between March and May 2023 by 14 teams, each comprising a leader, interviewers, and staff responsible for completing biomarker and performance tests. All team members underwent a 10-day training before data collection began.

HbA1c was tested using the A1CNow^R+^point-of-care testing device, which requires approximately 5 µL of blood sample and provides results within five minutes.^29^ The device features a built-in quality control function to ensure reliable results. It automatically detects issues such as insufficient or excessive blood volume, abnormal haemoglobin density, and extreme temperatures.^29^

### Measures

The primary outcomes were pre-T2D and UDD.

- Pre-T2D was defined using WHO’s HbA1c criteria (5.7% −6.4%). For the risk factor analysis, the comparison group included individuals without T2D or HbA1c <5.7%, while those diagnosed or undiagnosed with T2D were excluded.
- UDD was defined as an HbA1c level >6.5% without prior diagnosis by a healthcare professional. The same comparison group and exclusion criteria used for pre-T2D were used for risk factor analysis.

The explanatory variables included place of residence (urban or rural), sex (male or female), age (18-29, 30-39, 40-49, > 50 years), level of education (none, primary, secondary, high school, or higher), and wealth quintile, using Principal Component Analysis (ranging from poorest to wealthiest). The urban and rural classifications followed the criteria set by the National Institute of Statistics (NIS). We also considered factors such as obesity, physical activity, tobacco use, waist circumference, fruit and vegetable consumption, high blood pressure, total cholesterol, triglyceride, and alcohol consumption. Additional details on the measurement and definition of these variables are presented in Table 1.

**Table 1:** Category and measurement of metabolic risk factors.

| Variable | Category and measurement |
| --- | --- |
| Obesity | Underweight (BMI < 18.5), Normal weight ($18.5 \leq \text{BMI} \leq 25$), Overweight ($25 \leq \text{BMI} < 30$), Obese (BMI $\geq 30$)<sup>30</sup> |
| Physical activities | Weekly physical activity was assessed using Metabolic Equivalents of Task (MET) minutes.<sup>31-34</sup> Participants self-reported the frequency and duration of moderate- and vigorous-intensity physical activity at work and non-work. MET values were assigned (4 for moderate and 8 for vigorous), and the total weekly MET minutes were |
| | calculated.<sup>31-34</sup> Participants were then classified into four groups: < 300 MET-minutes, 300 to < 600 MET-minutes, 600 to < 1200 MET-minutes (recommended), and ≥ 1200 MET-minutes (additional benefits).<sup>31 33 34</sup> |
| Tobacco use | Participants were classified as current tobacco users (current smoking/smokeless use daily or less than daily), former users (previous smoking/smokeless use daily or less than daily, but not currently), and non-users. Electronic cigarette smokers were also included in this study. |
| Waist circumference | Normal (male: ≤ 102 cm, female: ≤ 88 cm), High (male: > 102 cm, female: > 88 cm) |
| Fruit and vegetable consumption | Fruit and vegetable consumption was classified as meeting WHO recommendations (≥ five servings per day or approximately 400 g of fruit and vegetables combined) or not (< 5 servings per day).<sup>35</sup> |
| High blood pressure | High blood pressure is classified as "Never," "Ever," and "Uncontrolled." Uncontrolled blood pressure is defined as an average systolic ≥ 140 mmHg or diastolic ≥ 90 mmHg from the last two of three measurements.<sup>36</sup> "Ever" refers to individuals diagnosed with hypertension but not classified as uncontrolled during the survey. "Never" denotes those not diagnosed with hypertension by health professionals or detected in the survey. |
| Total cholesterol level | Elevated (≥ 240 mg/dL)<sup>37</sup> |
| Triglyceride levels | Elevated (≥ 150 mg/dL)<sup>38</sup> |
| Alcohol consumption | Low risk, high risk (assessed using AUDIT-C) <sup>39</sup> |

#### Analysis

Both descriptive and inferential statistical methods were used for the analysis. Weighted results were used for univariate and bivariate analyses, whereas unweighted results were used for multivariate analyses. Records with missing HbA1c results were excluded, but missing values for explanatory variables were retained to preserve the sample size. The descriptive analysis employed sampling weights to match Cambodia’s age-sex population and urban/rural distribution, using proportions and frequencies for categorical variables and means and standard deviations for continuous variables. The bivariate analysis utilised Chi-square or Fisher’s exact tests for categorical variables related to prediabetes and undiagnosed diabetes. Variables with a P-value of 0.2 or lower were included in the multivariate analysis, using generalised linear models (GLM) with binomial specification and stepwise selection to find the best-fitting model based on the lowest Akaike information criterion (AIC). No collinearity was identified using the variance inflation factor (VIF) with a cutoff value of 5 VIF. However, we identified the interaction between sex and age, pre-T2D status, and UDD status. This was addressed by combining sex and age into a single variable. Analysis and reporting were conducted using the gt_summary and MASS packages in R with a significance threshold of p < 0.05.

### Ethical Considerations

The National Ethics Committee for Health Research approved the World Health Survey Plus protocol in Cambodia on 20 July 2022 (approval code: 221; NECHR). Written informed consent was obtained from all the participants during the initial stages. Owing to concerns regarding the potential misuse of their signatures among participants, we subsequently requested the NECHR to transition to verbal consent.

#### Patients and public involvement

None of the participants was involved in developing the research questions, outcome measures, study design, or recruitment.

## RESULTS

### Participant characteristics

#### Socio-demographic characteristics

As shown in Table 2, this study included 5,271 participants, most of whom were female (52.2%) and lived in rural areas (58.2%). The average age was 40.8 years, with the most significant proportion in the 18-29 age group (29.6%). Most participants were married or living together (79.7%) and had low educational attainment, with 67.3% having completed primary school or less.

**Table 2.** Socio-demographic characteristics of participants.

| **Characteristic** | **Unweighted** | **Weighted** |  |
| --- | --- | --- | --- |
|  | **N = 5,271** | **N= 5271** | **95% CI** |
| Type of community |  |  |  |
| Rural | 3,214 (61.0) | 58.2 | 56, 61 |
| Urban | 2,057 (39.0) | 41.8 | 39, 44 |
| Sex of participant |  |  |  |
| Male | 1,616 (30.7) | 47.8 | 45, 50 |
| Female | 3,655 (69.3) | 52.2 | 50, 55 |
| Age in years [mean] | 47.3 (14.9) | 40.8 | 40, 41 |
| Age group (years) |  |  |  |
| 18-29 | 669 (12.7) | 29.6 | 27, 32 |
| 30-39 | 1,114 (21.1) | 24.6 | 23, 27 |
| 40-49 | 1,087 (20.6) | 17.4 | 16, 19 |
| 50+ | 2,401 (45.6) | 28.4 | 26, 30 |
| Marital status |  |  |  |
| Married or living together | 4,086 (77.5) | 79.7 | 78, 82 |
| Never married | 297 (5.6) | 11.6 | 10, 13 |
| Separated/divorced/windowed | 888 (16.8) | 8.7 | 7.6, 10 |
| Socio-economic class |  |  |  |
| Poorest | 1,062 (20.1) | 14.3 | 13, 16 |
| Poor | 1,062 (20.1) | 17.1 | 15, 19 |
| Middle | 1,060 (20.1) | 20.1 | 18, 22 |
| Rich | 1,053 (20.0) | 20.8 | 19, 23 |
| Richest | 1,034 (19.6) | 27.8 | 26, 30 |
| Education level |  |  |  |
| Never attended school | 1,143 (21.7) | 14.5 | 13, 16 |
| Less than primary school | 1,830 (34.7) | 25.9 | 24, 28 |
| Primary school completed | 1,213 (23.0) | 26.9 | 25, 29 |
| Secondary school completed | 640 (12.1) | 18.2 | 16, 20 |
| High school completed | 301 (5.7) | 9.5 | 8.1, 11 |
| College/pre-university/University completed | 144 (2.7) | 5.1 | 4.1, 6.3 |

*Abbreviation: CI, confident interval*

#### Modifiable risk characteristics

More than half of the participants (53%) were classified as physically inactive, and 18.1% were current tobacco users (Table 3). A quarter (24.7%) of the participants were at high risk of alcohol use disorders. Approximately 57% of the participants consumed fewer than five servings of fruits and vegetables daily, with an average of 2.0 servings of fruits and 2.4 of vegetables. The average BMI was 21.4; 23.3% were overweight, 13.4% were obese, and 9.9% had high waist circumference. Elevated triglyceride and total cholesterol levels were found in 40.8% and 11.0% of the participants, respectively. Uncontrolled hypertension was present in 7.0% of participants, whereas 14.2% had previously been diagnosed with hypertension.

**Table 3.**
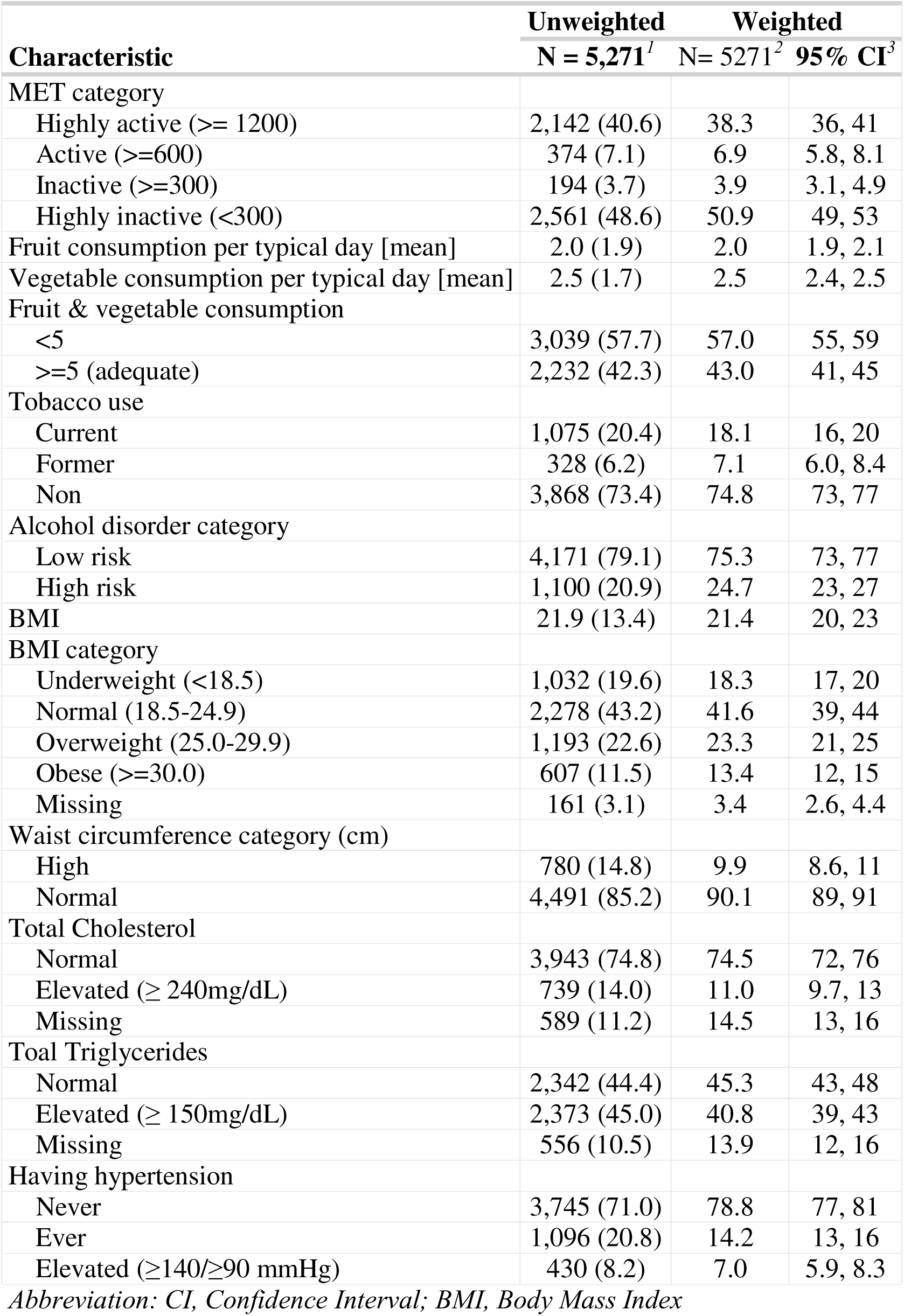
Modifiable risk characteristics of participants.

| Characteristic | Unweighted | Weighted |  |
| --- | --- | --- | --- |
|  | N = 5,271<sup>1</sup> | N = 5271<sup>2</sup> | 95% CI<sup>3</sup> |
| MET category |  |  |  |
| Highly active ($\geq 1200$) | 2,142 (40.6) | 38.3 | 36, 41 |
| Active ($\geq 600$) | 374 (7.1) | 6.9 | 5.8, 8.1 |
| Inactive ($\geq 300$) | 194 (3.7) | 3.9 | 3.1, 4.9 |
| Highly inactive ($< 300$) | 2,561 (48.6) | 50.9 | 49, 53 |
| Fruit consumption per typical day [mean] | 2.0 (1.9) | 2.0 | 1.9, 2.1 |
| Vegetable consumption per typical day [mean] | 2.5 (1.7) | 2.5 | 2.4, 2.5 |
| Fruit & vegetable consumption |  |  |  |
| <5 | 3,039 (57.7) | 57.0 | 55, 59 |
| $\geq 5$ (adequate) | 2,232 (42.3) | 43.0 | 41, 45 |
| Tobacco use |  |  |  |
| Current | 1,075 (20.4) | 18.1 | 16, 20 |
| Former | 328 (6.2) | 7.1 | 6.0, 8.4 |
| Non | 3,868 (73.4) | 74.8 | 73, 77 |
| Alcohol disorder category |  |  |  |
| Low risk | 4,171 (79.1) | 75.3 | 73, 77 |
| High risk | 1,100 (20.9) | 24.7 | 23, 27 |
| BMI | 21.9 (13.4) | 21.4 | 20, 23 |
| BMI category |  |  |  |
| Underweight ($< 18.5$) | 1,032 (19.6) | 18.3 | 17, 20 |
| Normal (18.5-24.9) | 2,278 (43.2) | 41.6 | 39, 44 |
| Overweight (25.0-29.9) | 1,193 (22.6) | 23.3 | 21, 25 |
| Obese ($\geq 30.0$) | 607 (11.5) | 13.4 | 12, 15 |
| Missing | 161 (3.1) | 3.4 | 2.6, 4.4 |
| Waist circumference category (cm) |  |  |  |
| High | 780 (14.8) | 9.9 | 8.6, 11 |
| Normal | 4,491 (85.2) | 90.1 | 89, 91 |
| Total Cholesterol |  |  |  |
| Normal | 3,943 (74.8) | 74.5 | 72, 76 |
| Elevated ($\geq 240$mg/dL) | 739 (14.0) | 11.0 | 9.7, 13 |
| Missing | 589 (11.2) | 14.5 | 13, 16 |
| Total Triglycerides |  |  |  |
| Normal | 2,342 (44.4) | 45.3 | 43, 48 |
| Elevated ($\geq 150$mg/dL) | 2,373 (45.0) | 40.8 | 39, 43 |
| Missing | 556 (10.5) | 13.9 | 12, 16 |
| Having hypertension |  |  |  |
| Never | 3,745 (71.0) | 78.8 | 77, 81 |
| Ever | 1,096 (20.8) | 14.2 | 13, 16 |
| Elevated ($\geq 140/\geq 90$ mmHg) | 430 (8.2) | 7.0 | 5.9, 8.3 |

Abbreviation: CI, Confidence Interval; BMI, Body Mass Index

#### Prevalence of pre-T2D and undiagnosed diabetes

Of the 41,04 individuals included, the weighted prevalence of pre-T2D and UDD was 27.2% and 10.9%, respectively, with a prevalence of diagnosed T2D of 7.3%.

#### Bivariate analysis of factors associated with pre-T2D and UDD

Bivariate analysis identified eight potential variables associated with pre-T2D and/or UDD (Supplementary Table 1). A higher prevalence of pre-T2D and UDD is associated with older age, living in urban areas, higher BMI, higher waist circumference, elevated blood pressure, and elevated triglyceride levels. Male sex was significantly associated with a high prevalence of pre-T2D only, whereas elevated total cholesterol level was associated with a higher prevalence of UDD only.

#### Multivariable logistic regression of factors associated with pre-T2D and UDD

In multivariate analysis, six variables were independently associated with a high prevalence of pre-T2D and/or UDD and retained in the model (Table 4).

**Table 4.**
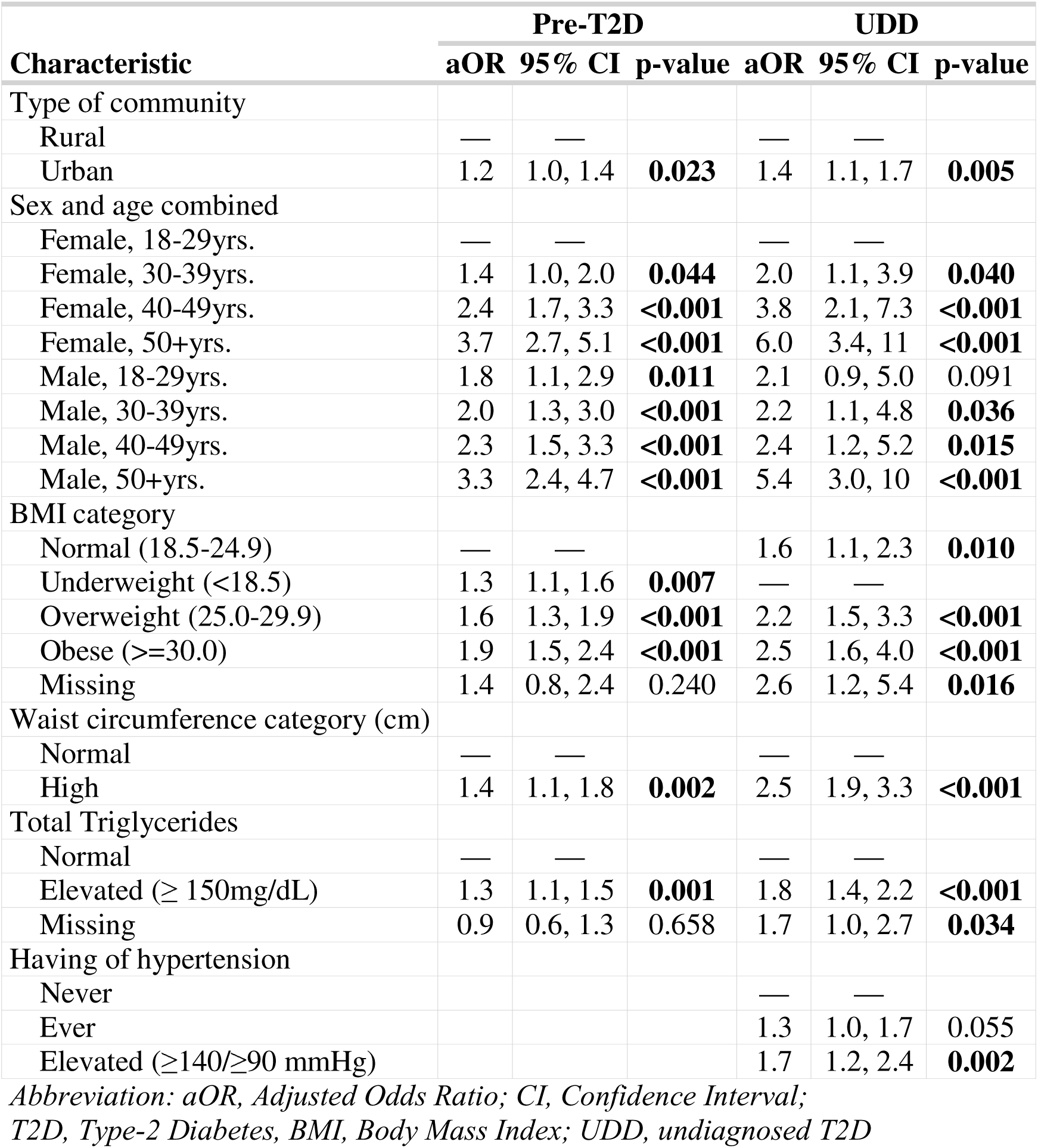
Independent factors associated with pre-T2D and UDD.

#### Factors independently associated with pre-T2D

Higher odds of pre-T2D were observed in individuals residing in urban areas (aOR = 1.2, 95% CI: 1.0 - 1.4), females aged 30-39 (aOR = 1.4, 95% CI: 1.0 - 2.0), females aged 40-49 (aOR = 2.4, 95% CI: 1.7 - 3.3), females aged 50+ (aOR = 3.8, 95% CI: 2.8 - 5.1), and males aged 30-39 (aOR = 2.0, 95% CI: 1.3 - 3.0), males aged 40-49 (aOR = 2.3, 95% CI: 1.5 - 3.3), males aged 50+ years group compared to the 18-29 years group (aOR = 3.3, 95% CI: 2.4 - 4.7) relative to females aged 18-29. Higher odds were also observed in individuals categorised as overweight (aOR = 1.6, 95% CI: 1.3 - 1.9), obese (aOR = 1.9, 95% CI: 1.5 – 2.4), with a high waist circumference (aOR = 1.4, 95% CI: 1.1 - 1.8), and with elevated total triglycerides (aOR = 1.3, 95% CI: 1.1 - 1.5).

#### Factors independently associated with UDD

Higher odds of UDD were observed in urban residents (aOR = 1.4, 95% CI: 1.1 - 1.7) and females aged 30-39 (aOR = 2.0, 95% CI: 1.1 - 3.9), females aged 40-49 (aOR = 3.8, 95% CI: 2.1 - 7.3), females aged 50+ (aOR = 6.0, 95% CI: 3.4 – 11.0), and males aged 30-39 (aOR = 2.2, 95% CI: 1.1 - 4.8), males aged 40-49 (aOR = 2.4, 95% CI: 1.2 - 5.2), and males aged 50+ years (aOR = 5.4, 95% CI: 3.0, 10.0), compared to females in the 18-29 year.

UDD was also more likely in individuals who were overweight (aOR = 2.2, 95% CI: 1.5 - 3.3), obese (aOR = 2.5, 95% CI: 1.6 - 4.0), had high waist circumference (aOR = 2.5, 95% CI: 1.9 - 3.3), elevated total triglyceride levels (aOR = 1.8, 95% CI: 1.4 - 2.2), or elevated blood pressure (aOR = 1.2, 95% CI: 1.2 - 2.4).

## DISCUSSION

Our investigation showed a high prevalence of pre-T2D and UDD among adults aged 18 years or older, at 27.3% and 10.9%, respectively. The combined prevalence of pre-T2D, diagnosed, and UDD is approximately 45%, indicating the substantial burden of pre-T2D and T2D in Cambodia. This study used HbA1c to estimate pre-T2D prevalence in Cambodia. Comparing our prevalence to the global prevalence presents challenges owing to variations in diagnostic tools and cutoff criteria. Studies using HbA1c have reported a higher prevalence than those using IGT or IFG.^5–7^ ^40^ ^41^ Our pre-T2D prevalence was approximately threefold higher than the globally estimated 10% using IFG.[3] Regional comparisons also proved challenging because of the different age ranges of the study population. However, when considering only HbA1c studies, our pre-T2D prevalence was lower than 34.6% in Vietnam (2015, ≥20 years) ^8^, higher than 14.2% in Malaysia (2006-2012, ≥35 years) ^42^, and 21.7% in China (2010, ≥18 years) ^43^.^43^. Our pre-T2D prevalence was lower than that in some European studies, including 24.7% in Swiss adults (25-41 years) ^5^, approximately 30% in Spanish adults (≥18 years) ^9^, and 42% in the Caribbean’s Barbados (≥25 years) ^7^. These results revealed a significant prevalence of potential future T2D in Cambodia, highlighting the urgent need for public health measures to address this concern.

We found a 10.9% prevalence of UDD, comprising approximately 60% of all T2D cases in our study (detected by HbA1c or self-reported). This proportion is slightly higher than the 55.2% previously reported for Cambodia by *Te et al*.^16^ and markedly higher than the estimates for the Southeast Asian region at 51.3% ^44^, suggesting potential gaps in screening, healthcare access, and disease awareness.

Advanced age has emerged as a significant risk factor for both pre-T2D and undiagnosed diabetes, with the likelihood increasing markedly with age. This aligns with prior research identifying age as a non-modifiable risk factor for T2D. ^7^ ^17^ ^18^ ^20^ ^26^ ^45–49^ Biological mechanisms include increasing insulin resistance and pancreatic beta-cell dysfunction over time, which contribute to elevated blood sugar levels and T2D development.^50^ Interestingly, we observed an interaction between age and sex. Males exhibited a higher prevalence of pre-T2D and developed the condition earlier, whereas females surpassed males in terms of prevalence at 40 years and beyond, possibly owing to their inherent biological advantages. Older females are likely to encounter more significant behavioural risks than males.^50^ These results indicate that interventions should be tailored based on age and sex to maximise effectiveness.

Urban residence was associated with higher odds of developing both pre-T2D and UDD, likely because of urban lifestyle and environmental factors. A prior study in Cambodia reported a higher prevalence of overweight and obesity in urban populations.^51^ Urban settings often promote dietary shifts towards processed foods, increased sedentary behaviour, or reduced physical activity, all of which contribute to metabolic risk.

Our study also found that pre-T2D and UDD prevalence were strongly associated with overweight and obesity, high waist circumference, elevated triglyceride levels, and elevated blood pressure. This finding is consistent with previous findings that obesity and high waist circumference are independent risk factors associated with T2D. ^50^ ^52^ ^53^ Individuals who are overweight or obese and have a high waist circumference tend to accumulate more visceral fat, impairing insulin sensitivity and increasing the risk of T2D. Similarly, elevated triglyceride and blood pressure levels are associated with insulin resistance, a key factor in T2D development.^50 52–54^

The literature shows that lifestyle factors, such as lack of physical activity, low fruit and vegetable consumption, alcohol consumption, and tobacco use, are linked to pre-T2D and T2D.^55–57^ These findings are mainly from prospective cohort studies.^55–57^ However, our study did not find significant associations between these variables in bivariate and adjusted logistic regressions. This discrepancy may stem from the self-reported biases in our cross-sectional design. Future studies with longitudinal designs could better capture the long-term impacts of lifestyle factors on T2D risk.

### Implications of the study

Early detection and intervention of pre-T2D through lifestyle changes or medication can reduce the risk of progression by up to 49% and prevent more severe T2D. ^30^ ^58^ The study highlights the importance of enhanced prevention and T2D screening, especially for higher-risk groups. Successful lifestyle interventions have been seen in various countries, including the U.S., Finland, Japan, India, and China.^59–62^ Although this study did not demonstrate significant associations, increasing access to healthy food and more stringent regulation of unhealthy products can also have (indirect) effects in the long term.^59–64^

### Strengths and limitations

A key strength of our study is its large, nationally representative sample size and the use of HbA1c for diagnosing pre-T2D and T2D, which provides a reliable assessment of the glycaemic status. However, the limitations include the cross-sectional design, which limits causal inference and potential reporting biases in self-reported lifestyle factors. Additionally, while HbA1c is a widely accepted diagnostic tool, variations in cutoff values across studies may affect prevalence comparisons. Future research should consider longitudinal approaches to understand risk trajectories and intervention effectiveness better.

## CONCLUSION

Our study revealed a high prevalence of pre-T2D and UDD in Cambodia, underscoring the critical need for targeted public health intervention. Significant risk factors included urban residence, advanced age, excess body weight, obesity, increased waist circumference, elevated blood pressure, and high triglyceride levels. Prevention and screening efforts should prioritise these high-risk populations. A comprehensive multi-sectoral approach encompassing healthcare services, community-based initiatives, and health promotion strategies is imperative to mitigate these risks and enhance diabetes management in Cambodia.

## Data Availability

All data produced in the present study are available upon reasonable request to the authors.

https://apps.who.int/healthinfo/systems/surveydata/index.php/collections/central

## ACKNOWLEDGEMENTS

We thank all participants and data collectors for meticulously adhering to the study instructions. Our appreciation also goes to WHO and HelpAge International, mainly thanks to Dr. Nirmala Naidoo from the WHO’s Department of Health Statistics and Information Systems in Geneva, Switzerland, for providing exceptional technical assistance throughout the WHS + design and data collection stages.

## AUTHOR CONTRIBUTIONS

CC, HS, SC, and PK designed the study; SC analysed the data; SC wrote the first draft; SC, PK, CN, SL, PW, KTM, GMK, KKG, NN, PI, CC, and HS provided subsequent inputs; and all authors commented on and approved the final version of the manuscript. The SC is responsible for the overall content [as a guarantor].

## DATA AVAILABILITY STATEMENT

Data are available upon reasonable request through https://apps.who.int/healthinfo/systems/surveydata/index.php/collections/central.

