## Supplementary figures and images for "Prevalence and factors associated with prediabetes and undiagnosed diabetes in Cambodia: Cross-sectional study based on the World Health Survey Plus 2023"

### Supplementary Figure 1

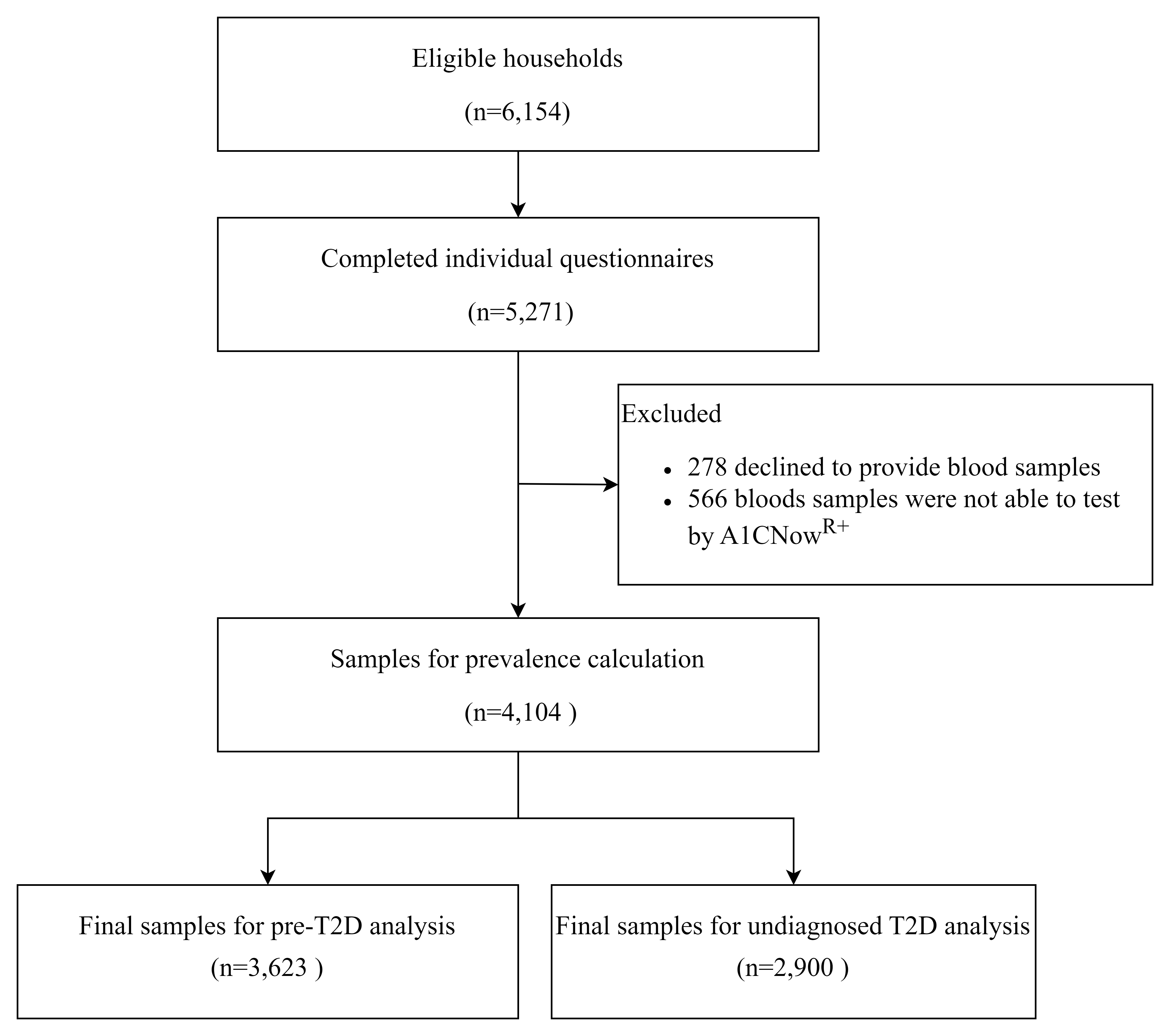
