## Supplementary Table 1 for "Prevalence and factors associated with prediabetes and undiagnosed diabetes in Cambodia: Cross-sectional study based on the World Health Survey Plus 2023"

Supplementary Table 1. Bivariate analysis results of the association between potential factors and pre-T2D and undiagnosed T2D

|  | **Pre-T2D** | | | **UDD** | | |
| --- | --- | --- | --- | --- | --- | --- |
| **Characteristic** | **No diabetes**, N = 2419 | **Pre-T2D**, N = 1204 | **p-value** | **No diabetes**, N = 2419 | **Undiagnosed T2D**, N = 481 | **p-value** |
| Type of community |  |  | **<0.001** |  |  | **<0.001** |
| Rural | 1,611 (72.8) | 726 (27.2) |  | 1,611 (89.9) | 257 (10.1) |  |
| Urban | 808 (62.8) | 478 (37.2) |  | 808 (80.3) | 224 (19.7) |  |
| Sex of participant |  |  | **<0.001** |  |  | >0.9 |
| Male | 725 (64.5) | 395 (35.5) |  | 725 (86.2) | 141 (13.8) |  |
| Female | 1,694 (73.2) | 809 (26.8) |  | 1,694 (86.4) | 340 (13.6) |  |
| Age group (years) |  |  | **<0.001** |  |  | **<0.001** |
| 18-29 | 446 (81.8) | 101 (18.2) |  | 446 (94.4) | 23 (5.6) |  |
| 30-39 | 627 (69.6) | 207 (30.4) |  | 627 (89.6) | 65 (10.4) |  |
| 40-49 | 516 (62.4) | 261 (37.6) |  | 516 (81.5) | 99 (18.5) |  |
| 50+ | 830 (55.7) | 635 (44.3) |  | 830 (73.7) | 294 (26.3) |  |
| MET category |  |  | 0.4 |  |  | 0.6 |
| Highly active (>= 1200) | 1,026 (68.6) | 499 (31.4) |  | 1,026 (86.4) | 192 (13.6) |  |
| Active (>=600) | 177 (69.8) | 83 (30.2) |  | 177 (82.2) | 38 (17.8) |  |
| Inactive (>=300) | 90 (79.7) | 51 (20.3) |  | 90 (90.5) | 20 (9.5) |  |
| Highly inactive (<300) | 1,126 (68.3) | 571 (31.7) |  | 1,126 (86.5) | 231 (13.5) |  |
| Fruit and vegetable consumption |  |  | 0.3 |  |  | >0.9 |
| <5 | 1,384 (70.3) | 666 (29.7) |  | 1,384 (86.4) | 274 (13.6) |  |
| >=5 (adequate) | 1,035 (67.3) | 538 (32.7) |  | 1,035 (86.2) | 207 (13.8) |  |
| Smoking |  |  | 0.051 |  |  | 0.061 |
| Current | 476 (65.3) | 280 (34.7) |  | 476 (88.4) | 93 (11.6) |  |
| Former | 129 (59.9) | 78 (40.1) |  | 129 (77.2) | 39 (22.8) |  |
| Non | 1,814 (70.7) | 846 (29.3) |  | 1,814 (86.6) | 349 (13.4) |  |
| Alcohol disorder category |  |  | 0.8 |  |  | 0.5 |
| Low risk | 1,909 (68.8) | 965 (31.2) |  | 1,909 (86.7) | 384 (13.3) |  |
| High risk | 510 (69.6) | 239 (30.4) |  | 510 (85.1) | 97 (14.9) |  |
| BMI category |  |  | **<0.001** |  |  | **<0.001** |
| Underweight (<18.5) | 421 (75.5) | 213 (24.5) |  | 421 (91.5) | 42 (8.5) |  |
| Normal (18.5-24.9) | 1,245 (77.6) | 461 (22.4) |  | 1,245 (89.3) | 198 (10.7) |  |
| Overweight (25.0-29.9) | 509 (61.1) | 334 (38.9) |  | 509 (82.6) | 139 (17.4) |  |
| Obese (>=30.0) | 206 (49.7) | 175 (50.3) |  | 206 (75.1) | 90 (24.9) |  |
| Missing | 38 (51.6) | 21 (48.4) |  | 38 (77.4) | 12 (22.6) |  |
| Waist circumference category (cm) |  |  | **0.005** |  |  | **<0.001** |
| High | 232 (57.6) | 207 (42.4) |  | 232 (64.8) | 143 (35.2) |  |
| Normal | 2,187 (70.1) | 997 (29.9) |  | 2,187 (88.7) | 338 (11.3) |  |
| Total Cholesterol |  |  | 0.11 |  |  | **<0.001** |
| Normal | 2,009 (69.9) | 949 (30.1) |  | 2,009 (88.5) | 360 (11.5) |  |
| Elevated (≥ 240mg/dL) | 272 (60.7) | 199 (39.3) |  | 272 (69.0) | 101 (31.0) |  |
| Missing | 138 (70.2) | 56 (29.8) |  | 138 (88.0) | 20 (12.0) |  |
| Total Triglycerides |  |  | **<0.001** |  |  | **<0.001** |
| Normal | 1,349 (75.7) | 525 (24.3) |  | 1,349 (92.3) | 153 (7.7) |  |
| Elevated (≥ 150mg/dL) | 966 (61.4) | 632 (38.6) |  | 966 (80.0) | 300 (20.0) |  |
| Missing | 104 (62.1) | 47 (37.9) |  | 104 (73.5) | 28 (26.5) |  |
| Having hypertension |  |  | **<0.001** |  |  | **<0.001** |
| Never | 1,912 (71.3) | 824 (28.7) |  | 1,912 (89.3) | 277 (10.7) |  |
| Ever | 356 (65.4) | 268 (34.6) |  | 356 (76.3) | 130 (23.7) |  |
| Elevated (≥140/≥90 mmHg) | 151 (47.5) | 112 (52.5) |  | 151 (66.9) | 74 (33.1) |  |
| Abbreviation: MET, Metabolic Equivalent of Task; BMI, Body Mass Index; UDD, undiagnosed T2D | | | | | | |
